# Subthalamic Nucleus Deep Brain Stimulation Modulates Strabismus in Parkinson’s Disease

**DOI:** 10.1101/2024.11.07.24316853

**Authors:** Palak Gupta, Sinem Balta Beylergil, Camilla Kilbane, Cameron C. McIntyre, Angela M Noecker, Aasef G. Shaikh, Fatema F. Ghasia

## Abstract

**Objective:** Parkinson’s disease (PD) is a neurodegenerative disorder characterized by motor and non-motor symptoms. Visual impairments, such as strabismus (misalignment of the eyes during gaze holding), affect up to two-thirds of PD patients, impacting their quality of life. Conventional treatments offer limited relief, prompting exploration of alternatives like deep brain stimulation (DBS) of subthalamic nucleus (STN). This pilot study aims to assess whether STN DBS can alleviate PD-related strabismus and identify specific STN regions associated with favorable outcomes. We hypothesize that STN DBS improves strabismus by modulating subthalamic connectivity with the cerebellum, hence volume of activate tissue (VTA) generated with DBS will be in dorsal STN.

**Methods:** We studied 12 PD patients with bilateral STN DBS and five healthy controls. Clinical assessments, eye movement measurements using high-resolution eye tracking, and patient-specific DBS models were employed. Analysis included the VTA models, revealing distinct effects based on the location within the STN.

**Results:** We found significant strabismus in 66% of PD patients. STN DBS improved strabismus in 75% of cases. The improvement was associated with dorsal STN stimulation. STN DBS exacerbated strabismus in 25% of PD patients. The VTA in these participants were located in the ventral aspect of the STN.

**Discussion:** The findings highlight the significant effects of STN DBS on strabismus in PD, further offering insights into the complex interplay between neurodegeneration and control of eye alignment. This approach, combining clinical assessments, advanced eye tracking, and DBS computational modeling, contributes valuable knowledge towards targeted interventions for visual impairments in PD.

## Introduction

Parkinson’s disease (PD) is a neurodegenerative condition that affects approximately 10 million people worldwide, leading to a range of motor and non-motor symptoms. Visual problems in PD are multifaceted and more common than appreciated. They encompass issues such as blepharospasm, dry eyes, reduced blinking, visual hallucinations, decreased visual acuity, and contrast sensitivity [1-5]. The eye movement abnormalities are not uncommon in PD, manifesting as abnormal rapid gaze shifts, saccades, often accompanied by increased saccadic intrusions, which can make daily tasks such as reading and scanning the visual scene difficult [1, 6-10].

Vergence insufficiency, the inability of the two eyes to coordinate in response to shifting the gaze from far to near (convergence) or vice versa (divergence), is common in PD [3, 7, 9-11]. Strabismus, the misalignment of the eyes, is also common in PD. Strabismus can be attributed to both, vergence insufficiency as well as subcortical deficit specific to PD and can cause double vision in about one-third of PD patients [1, 4, 5, 9, 10, 12-15]. Strabismus and vergence insufficiency are even more evident on instrumented measures, as noted in our previous study where 55% of patients had increased eye misalignment under binocular viewing condition (i.e., tropia). Nearly all participants had impaired alignment under monocular viewing (i.e., phoria), and had vergence insufficiency [12]. The abnormal binocular control translates in lower visual health in PD with significant reductions in visual quality of life survey scores, particularly for near vision [1, 4, 14, 16-19]. The impact of these visual deficits extends beyond mere discomfort, significantly contributing to the higher prevalence of depression and anxiety commonly observed in PD [20, 21]. Collectively, these reports highlight the imperative need for effective interventions.

The visuomotor deficits such as vergence and strabismus respond poorly to conventional pharmacotherapy with levodopa[22, 23]. The fluctuation in convergence ability throughout the day poses a significant challenge in the ophthalmic management of PD patients, significantly affecting their quality of life. Progressive neurodegeneration in PD makes these patients unsuitable for ophthalmological interventions such as strabismus surgery to treat binocular disparity and double vision. The state-of-art surgical therapy with subthalamic nucleus region (STN) deep brain stimulation (DBS) frequently improves motor symptoms such as tremor, bradykinesia, hypokinesia, and rigidity, but its effects on binocular control leading to strabismus and related impairment in the quality of life are not yet known [1, 24, 25].

We present a pilot investigation where the objective was to demonstrate the strabismus phenomenology and provide insights into its potential treatment with STN DBS. We address two unanswered questions:

1. Can STN-DBS alleviate binocular misalignment, a factor contributing to strabismus in PD?
2. Which STN regions are linked to enhanced strabismus improvement while maintaining positive effects on motor symptoms?

These questions are motivated by the hypothesis that DBS of the dorsal STN improves strabismus in PD. The hypothesis is based on the premise that the dorsal STN communicates with the pre-cerebellar pontine neurons influencing the cerebellar outflow. The cerebellar outflow modulates the supra-oculomotor area (SOA) within the brainstem, the region where the strabismus angle sensitive neurons are located [26, 27]. Thus, STN DBS may treat strabismus in PD by modulating the cerebellar outflow that projects to SOA.

## Methods

### Study Participants & Clinical Assessments

We studied 12 PD (age: 66.58±8.15) and five healthy (age: 67±9.88) participants. The PD participants met the UK Brain Bank criteria for the diagnosis, their motor symptoms robustly responded to levodopa, and all PD participants were implanted with bilateral STN DBS. The measurements of motor symptoms, and eye movements were made when the STN DBS was on and when it was turned off. The visual acuity and stereo-acuity, refraction, and strabismus angle measurements at distance and near the time of eye movement recordings were noted. The presence of strabismus was assessed with a prism and alternate cover test in the appropriate diagnostic fields of gaze at 6 meters as well as at one-third of a meter in the primary position. We used the standard guidelines recommended by the pediatric ophthalmology and strabismus subsection of the American Academy of Ophthalmology preferred practice pattern to assess strabismus. Near point of convergence (NPC) was measured with a Prince ruler. Out of the 12 PD patients, only one patient did not have clinically measurable exodeviation at near. Only one control subject had a small intermittent exodeviation of 2 prism diopters at near, whereas the remaining age-matched controls did not have any clinically measured exodeviation at near. We defined normal intact vergence as an NPC of less than 10 cm. Out of the five healthy age-matched controls, all had intact vergence whereas one control had mild increase in NPC at 14 cm. In our study, all PD patients exhibited increased NPC (>10 cm). Demographic information, clinically measured motor features of PD – Unified Parkinson’s Disease Rating Scale (UPDRS-III), were recorded for every patient (**Table 1**). The research protocol was approved by The Department of Veterans Affairs, The University Hospitals, and The Cleveland Clinic Institutional Review Boards.

**Table 1:** Demographics, clinical features, and subthalamic deep brain stimulation parameters. ID: patient ID, yrs: years, UPDRS-III Unified Parkinson’s Disease Rating Scale part III, mg: milligram, V: volts, Hz: Herz, STN DBS: Subthalamic nucleus deep brain stimulation.

|  |  | STN DBS OFF |  |  | STN DBS ON |  |  |  |  |  | Left STN DBS parameters |  |  |  | Right STN DBS Parameters |  |  |  |
| --- | --- | --- | --- | --- | --- | --- | --- | --- | --- | --- | --- | --- | --- | --- | --- | --- | --- | --- |
| ID | Disease Duration (yrs) | UPDRS-III | Cover Test Distance | Cover Test Near | UPDRS-III | Cover Test Distance | Cover Test Near | Dopamine/day (mg) | DBS System | Lead Type | Active Contact | Voltage (V) | Pulse Width (microsecond) | Frequency (Hz) | Active Contact | Voltage (V) | Pulse Width (microsecond) | Frequency (Hz) |
| 1 | 11 | 51 | Ortho | 4X(T) | 6 | Ortho | 2X(T) | 1040 | Medtronic Activa PC | 3389 | 3 - C + | 2.5 | 60 | 160 | 3 - 2 + | 1.5 | 60 | 160 |
| 2 | 5 | 12 | Ortho | Ortho | 6 | Ortho | Ortho | 60 | Medtronic Activa PC | 3389 | 1 - 3 - 2 + | 3.3 | 60 | 130 | 2 - C + | 2.5 | 60 | 130 |
| 3 | 12 | 41 | Ortho | 12X(T) | 7 | Ortho | 2X(T) | 1505 | Medtronic Activa PC | 3389 | 1 - C + / 2 - C + | 3.6 | 90 | 125 | 1 - 2 - C + | 2.5 | 60 | 125 |
| 4 | 21 | 37 | 2RHT | 8X(T), 2RHT | 19 | 2RHT | 6X(T), 2RHT | 1100 | Medtronic Activa PC | 3389 | 2 - C + | 3.5 | 60 | 130 | 2 - C + | 3.5 | 60 | 130 |
| 5 | 9 | 48 | FlickX | 15X(T) | 12 | Ortho | 6X(T) | 400 | Medtronic Activa PC | 3389 | 1 - 3 - 2 + | 5 | 90 | 130 | 3 - C + | 3 | 90 | 130 |
| 6 | 10 | 42 | Ortho | 4X(T) | 23 |  |  | 300 | Medtronic Activa PC | 3389 | 2 - C + | 3.5 | 60 | 185 | 2 - 1 + | 4.4 | 60 | 185 |
| 7 | 18 | 76 | Ortho | 10X(T) |  | Ortho | 6X(T) | 500 | Medtronic Activa PC | 3389 | 2 - C + | 3 | 60 | 140 | 1 - 3 - C + | 3.5 | 60 | 140 |
| 8 | 14 | 41 | Ortho | 14X(T) | 16 | Ortho | 12X(T) | 610 | Medtronic<br>Activa PC | 3389 | 2 –<br>C + | 2.5 | 60 | 130 | 2 –<br>C + | 3 | 60 | 130 |
| 9 | 17 | 54 | Ortho | 9X(T) | 21 | Ortho | 6X(T) | 500 | Medtronic<br>Activa PC | 3389 | 3 –<br>C + | 3.5 | 60 | 180 | 1 –<br>2 –<br>C + | 2.3 | 60 | 180 |
| 10 | 8 | 25. | 2LHT | 6X(T),<br>2LHT | 9 | 3LHT | 8X(T),<br>3LHT | 500 | Abbott (St.<br>Jude<br>Medical)<br>Infinity | 6171 | 3 –<br>C + | 1 | 60 | 130 | 3 –<br>C + | 2.5 | 60 | 130 |
| 11 | 10 | 24 | FlickX | 4X(T) | 4 | Ortho | 4X(T) | 348 | Medtronic<br>Activa PC | 3389 | 1 –<br>3 + | 2.6 | 60 | 130 | 1 –<br>2 + | 1.3 | 60 | 130 |
| 12 | 10 | 30 | Ortho | 8X(T) | 16 | Ortho | 1X(T) | 600 | Medtronic<br>Activa PC | 3389 |  | 2.6 | 60 | 130 | 1 –<br>2 + | 1.3 | 60 | 130 |

### Objective Measures and Analysis of Eye Movements

A high-resolution eye tracker (EyeLink 1000plus) was used to measure binocular horizontal and vertical eye positions at 0.01° spatial and 500 Hz temporal resolution [13, 28, 29]. Strabismus was measured during monocular viewing (i.e., phoria) and binocular viewing (i.e., tropia). An infrared permissive filter blocked the visible light while allowing the non-viewing eye to be tracked with a video-oculography camera. The eye movements were measured when subjects focused the gaze on a visual target projected on a screen that was 55 cm away. The target was bright red in color on a white background and it subtended at 0.5° visual angle. Right and left eye movements were independently calibrated in monocular viewing condition. The experiment session that followed monocular calibration involved measurements in right eye viewing, left eye viewing and both eyes viewing, each session lasting 45 seconds. The detail of the infrastructure is outlined in our earlier publication [30].

The analysis was performed with a customized software program written in-house, as summarized in our recent study [12]. The key parameter of interest was the strabismus angle, which is the difference in vertical and horizontal positions of right and left eye, measured in monocular (phoria) and binocular (tropia) viewing conditions. We computed the composite difference between the right and left eye position using equation (1.1).

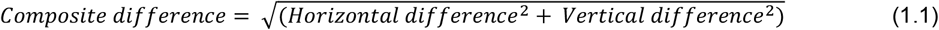

To objectively compare the strabismus angle across different participants, we parsed eye position signal within 2 millisecond epochs. Then we measured strabismus angles for each 2ms epochs. We then utilized the Empirical Cumulative Distribution Function (ECDF), a statistical tool to visualize and analyze the distribution of the strabismus angle dataset gathered from each 2ms epochs. The ECDF is a non-parametric estimator of the cumulative distribution function (CDF) associated with variables, in this case strabismus angles. It is calculated by first sorting the data in ascending order and then computing the cumulative sum of the sorted values. The resultant sequence of cumulative sums is then normalized by dividing each of the sum by total number of data points. The normalized cumulative sum represents the ECDF. The purpose of such analysis is to provide a visual representation of the data distribution without making assumptions for the underlying probability distribution.

In addition, we quantified strabismus angle over a 45 second timespan, computing the maximum eye position difference between the two eyes across the timespan, and the percent time during which the eyes were aligned maintaining fusion.

The statistical analysis was performed in MATLAB (MathWorks, Natick, MA) and SPSS. Age between controls and PD participants was compared with the Mann-Whitney U test and Kolmogorov–Smirnov (KS) test.

### Patient-specific DBS Models and Metrics

Our next analytical step involved determining the volume and location of the DBS electrical field within the STN region that corresponds to improvement, lack of change, or worsening of the strabismus angle. To conduct this analysis, we utilized StimVision, an academic DBS research software [8, 31-35]. Using the patients’ MRI data, StimVision generated a 3D representation of the DBS electrical field in each patient’s brain. This representation included the activated volumes (VTA) of the STN and subcortical brain regions surrounding the STN [8, 31-35]. The DBS models employed in this study were based on the clinically optimal stimulation settings in use at the time for each patient. These models provided various metrics, including VTA (mm^3^), the overlapping VTA and the STN volume (mm^3^), Euclidean distance between the centers of the STN and the distance between the center of the STN and the VTA along the dorso-ventral, antero-posterior, and medio-lateral axes.

## Results

The overarching goal of our study was to objectively analyze strabismus in PD, and to further examine the efficacy of STN DBS in its treatment. We also aimed to understand whether STN DBS parameters that improve strabismus also maintain their established effects on PD motor symptoms. Finally, we asked which STN regions are linked to enhanced strabismus improvement, while maintaining the effects on motor symptoms. For this study we recruited 12 PD participants and five healthy controls. The PD and healthy participants were age matched (PD: 66.58±8.15 years; healthy controls: 67±9.88 years, U=65; p=0.36). Mean duration since onset of symptoms was 12.06±4.6 years, while mean daily dopamine dose was 621.9±401.4 mg. All PD participants had improvement in motor symptoms with STN DBS as measured with UPDRS part III (UPDRS-III with STN DBS off=40.04±16.7, STN DBS on=12.63±6.68, t-test, p<0.001, **Table 1**). On clinical examination in DBS off state, the exotropia (outward deviation of one eye) on near vision was noted in all but one patient, who was well aligned (ortho-phoric). Two of the patients with exotropia also had hypertropia featuring vertical deviation. Clinical examination revealed improvement in strabismus with STN DBS, but the amount of improvement varied among participants.

**Figure 1** depicts an example of gaze holding during binocular viewing condition in one healthy participant (**Fig 1A**) and one PD participant with STN DBS off (**Fig 1B**) and on (**Fig 1C**). Both participants held gaze on a non-patterned visual target that was projected straight-ahead on the screen placed 55 cm from the subjects’ eyes. The vertical eye position is plotted on y-axis, while x-axis shows horizontal eye position. The right and left eyes of the healthy participant remained stable on straight-ahead orientation, at the intersection of dashed vertical and horizontal lines. Such binocular gaze holding behavior differed in the PD participant when the STN DBS was turned off as illustrated in **Fig 1B** (red trace shows right eye and orange trace left eye). The notable distinction in the PD participant compared to healthy control is that the left eye of the PD participant remained stable at the intersection of the gray horizontal and vertical lines, in straight-ahead orientation, but the right eye deviated to the right. At its worst the deviation was 7.2°, compared to the control value of 0.87° measured from the healthy participant. At its best, the eyes were precisely aligned, hence the binocular angle, the difference between the angular positions of the right and the left eye, varied within the range of 7.2° in PD with DBS off and 0.8° in healthy participant. Such eccentric drift of the right eye in right-ward direction is consistent with exotropia during binocular viewing condition. The right eye remained deviated for approximately 60% of the time (not evident from **Fig 1B**) and then it aligned again with the left eye, hence fusion was maintained only for 40% of the time as opposed to the healthy participant, whose the fusion was maintained for 100% of the time. The binocular alignment of this PD participant normalized when the DBS was turned on (overlapping blue and cyan traces at straight ahead, **Fig 1C**). The maximal deviation of the right eye in DBS on condition was 1.2°, while the binocular angle varied in the range of 1.2°. The fusion was maintained for 100% of the time in this condition.

**Figure 1:**
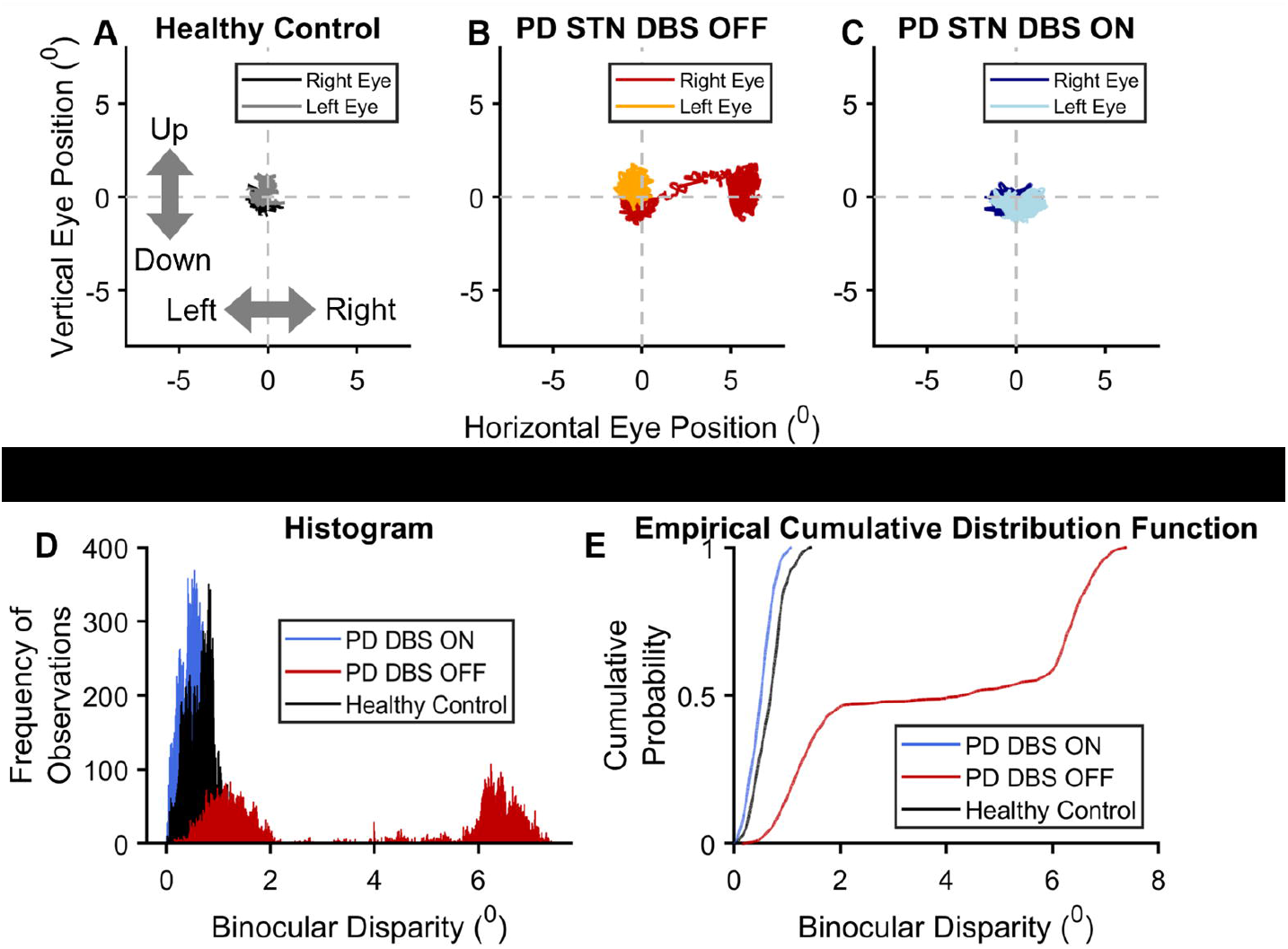
The effects of STN-DBS on strabismus in PD and its comparison with a healthy control. (A-C) The scatter plots show vertical eye position on the y-axis and horizontal eye position on the x-axis for a healthy control (A) and a PD participant with DBS off (B) and DBS on (C). Disparity in right eye position, seen as a shift to the right, when DBS was off in PD participant is corrected when it was turned on, and is maintained in a state that was comparable to the healthy control. Panels D depicts a summary of binocular disparity in the form of a histograms, providing a visual representation of the data. Panel E is an empirical cumulative distribution function, offering an additional way to understand the data distribution in PD with DBS on and off and its comparison with control.

The subsequent analysis parsed the right and left eye positions in 2 ms epochs. In each epoch we measured the binocular angle, depicting the severity of strabismus – larger angle correlating with more severe strabismus. Histograms in **Fig 1D** depict distributions of strabismus angles in healthy participants (black histograms), PD participant with DBS off (red histograms), and with DBS on (blue histogram). The histogram of strabismus angles is broadly distributed in PD participant with DBS off compared to DBS on. The latter’s distribution is comparable to that of the healthy control (**Fig 1D**). ECDFs were further compared between the DBS on and off states in the PD participant, in reference to the healthy control. The strabismus angles were sorted in the ascending order to compute the cumulative sum, and then normalized to form the ECDF. The normalized cumulative sum, depicting the cumulative probability is plotted on the y-axis of **Fig 1E**, while the x-axis displays the strabismus angle. The ECDF curve located on the left of the graph depicts minimal strabismus angles, but the increase in strabismus angles is associated with a shift of the ECDF curve to the right. As expected, the ECDF curve representing the healthy control is to the extreme left of the ECDF curve representing the PD participant with DBS off (red trace, **Fig 1E**). When the DBS was turned on, the ECDF of the PD participant shifted to the left (blue trace, **Fig 1E**), to the proximity of the healthy control’s ECDF (black trace, **Fig 1E**).

In addition to gaze holding during binocular viewing (i.e., tropia, an example is shown in **Figure 1**), we also measured strabismus, the deviation of the covered eye in monocular viewing condition (i.e., phoria). As expected, the PD participants showed superior control of binocular alignment and fusion maintenance in binocular viewing condition, compared to monocular viewing condition. During baseline (DBS off condition) the maximum strabismus angle (phoria) was consistently worse compared to binocular viewing condition (tropia) (**Supplementary Table s1**).

**Figure 2** depicts the summary of ECDF in PD compared to healthy controls. According to the effects of STN DBS, 12 PD participants were categorized into three groups: those showing no change (Group 1), those with improvement (Group 2), and those with worsening of strabismus (Group 3). Four PD participants were in Group 1, and had mild strabismus when DBS was off, and therefore had small trendwise improvement in strabismus in binocular (**Fig 2A**) and monocular viewing condition (**Fig 2E,I**) with STN DBS. There were six PD participants in Group 2, who had notably worse strabismus when STN DBS was off, as evidenced by the blue ECDF line positioned to the right of the black line in binocular (**Fig 2B,C**) and/or monocular (**Fig 2F,G,J,K**) viewing conditions. Three patients in Group 2 (i.e., Group 2A) had no strabismus in binocular viewing condition (**Fig 2B**), but the phoria was evident in the covered eye (**Fig 2F,J**). Three Group 2 patients (i.e., Group 2B) had strabismus in monocular and binocular viewing condition (**Fig 2C,G,K**). All six patients in Group 2 improved their strabismus when STN DBS was turned on, as can be seen from the ECDF shifting to the left, towards the control group. In Group 3 (n=2), similar to Group 2, the participants had robust strabismus, indicated by the blue line (ECDF during STN DBS off) positioned to the right of the black line (healthy control ECDF) in binocular (**Fig 2D**) and monocular viewing conditions (**Fig 2H,L**). STN DBS further exacerbated the strabismus in this group, as indicated by the red ECDF line, shifting further to the right (**Fig 2D,H,L**). **Table 2** presents a summary of the effects of STN DBS on both the maximum strabismus angle and the percentage of time fusion was sustained across three distinct groups under binocular and monocular viewing conditions. Notably, minor discrepancies in values were observed, resulting in a small effect size for Group 1. In Group 2A, a substantial effect size was evident during monocular conditions, while a smaller effect was noted in binocular viewing. Conversely, both monocular and binocular viewing conditions displayed notable effect sizes in Group 2B, highlighting the robust impact of STN DBS (**Table 2**). Contrastingly, Group 3 exhibited a decline in both the maximum strabismus angle and the percentage of time maintaining fusion (refer to **Table 2**). Despite differences in the effect size across the four categories, a statistically significant change associated with STN DBS was observed in the bin-wise measured binocular angle (ANOVA, p < 0.01).

**Table 2:**
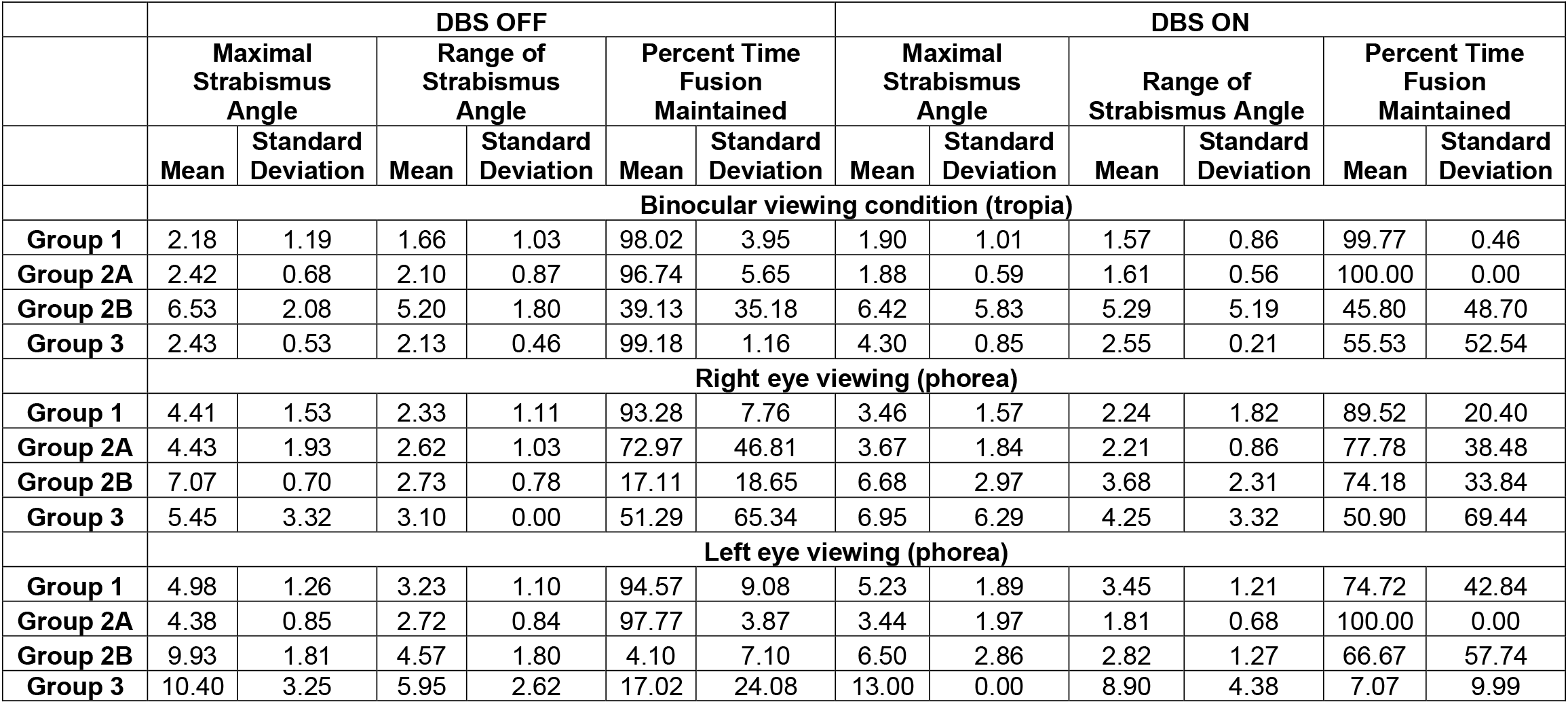
Visual angular canculation in response to STN DBS. Group 1: no response to STN DBS, baseline mild strabismus; Group 2A: baseline mild impairment of tropia but significant phorea, phorea improved by STN DBS; Group 2B: baseline significant tropia and phorea, both improved by STN DBS, Group 3: signficant phorea and tropia, both worsened with STN DBS.

|  | DBS OFF |  |  |  |  |  | DBS ON |  |  |  |  |  |
| --- | --- | --- | --- | --- | --- | --- | --- | --- | --- | --- | --- | --- |
|  | Maximal Strabismus Angle |  | Range of Strabismus Angle |  | Percent Time Fusion Maintained |  | Maximal Strabismus Angle |  | Range of Strabismus Angle |  | Percent Time Fusion Maintained |  |
|  | Mean | Standard Deviation | Mean | Standard Deviation | Mean | Standard Deviation | Mean | Standard Deviation | Mean | Standard Deviation | Mean | Standard Deviation |
|  | Binocular viewing condition (tropia) |  |  |  |  |  |  |  |  |  |  |  |
| **Group 1** | 2.18 | 1.19 | 1.66 | 1.03 | 98.02 | 3.95 | 1.90 | 1.01 | 1.57 | 0.86 | 99.77 | 0.46 |
| **Group 2A** | 2.42 | 0.68 | 2.10 | 0.87 | 96.74 | 5.65 | 1.88 | 0.59 | 1.61 | 0.56 | 100.00 | 0.00 |
| **Group 2B** | 6.53 | 2.08 | 5.20 | 1.80 | 39.13 | 35.18 | 6.42 | 5.83 | 5.29 | 5.19 | 45.80 | 48.70 |
| **Group 3** | 2.43 | 0.53 | 2.13 | 0.46 | 99.18 | 1.16 | 4.30 | 0.85 | 2.55 | 0.21 | 55.53 | 52.54 |
|  | Right eye viewing (phorea) |  |  |  |  |  |  |  |  |  |  |  |
| **Group 1** | 4.41 | 1.53 | 2.33 | 1.11 | 93.28 | 7.76 | 3.46 | 1.57 | 2.24 | 1.82 | 89.52 | 20.40 |
| **Group 2A** | 4.43 | 1.93 | 2.62 | 1.03 | 72.97 | 46.81 | 3.67 | 1.84 | 2.21 | 0.86 | 77.78 | 38.48 |
| **Group 2B** | 7.07 | 0.70 | 2.73 | 0.78 | 17.11 | 18.65 | 6.68 | 2.97 | 3.68 | 2.31 | 74.18 | 33.84 |
| **Group 3** | 5.45 | 3.32 | 3.10 | 0.00 | 51.29 | 65.34 | 6.95 | 6.29 | 4.25 | 3.32 | 50.90 | 69.44 |
|  | Left eye viewing (phorea) |  |  |  |  |  |  |  |  |  |  |  |
| **Group 1** | 4.98 | 1.26 | 3.23 | 1.10 | 94.57 | 9.08 | 5.23 | 1.89 | 3.45 | 1.21 | 74.72 | 42.84 |
| **Group 2A** | 4.38 | 0.85 | 2.72 | 0.84 | 97.77 | 3.87 | 3.44 | 1.97 | 1.81 | 0.68 | 100.00 | 0.00 |
| **Group 2B** | 9.93 | 1.81 | 4.57 | 1.80 | 4.10 | 7.10 | 6.50 | 2.86 | 2.82 | 1.27 | 66.67 | 57.74 |
| **Group 3** | 10.40 | 3.25 | 5.95 | 2.62 | 17.02 | 24.08 | 13.00 | 0.00 | 8.90 | 4.38 | 7.07 | 9.99 |

**Figure 2:**
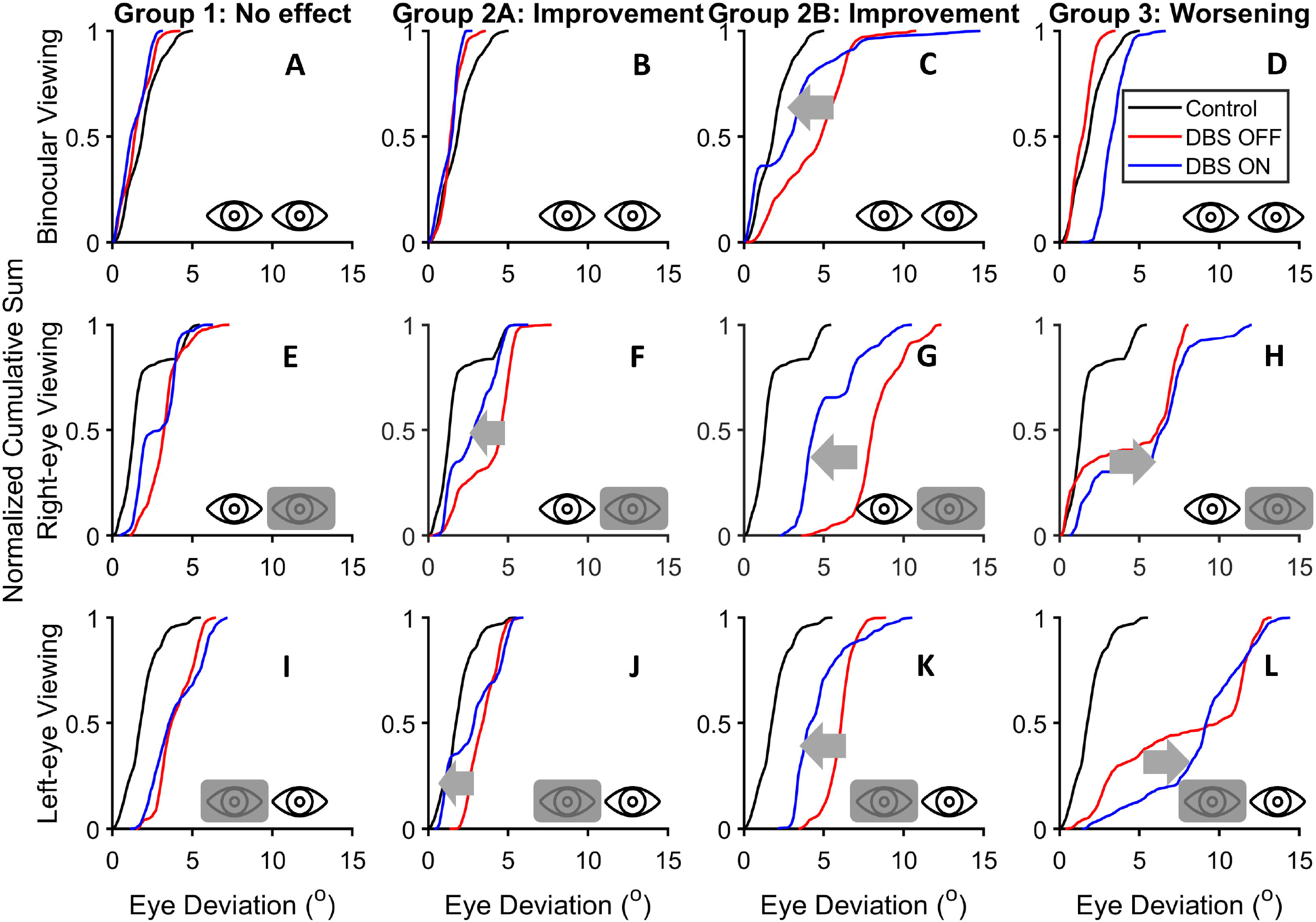
Summary of the STN-DBS effects on strabismus in 12 PD participants. In each plot the x-axis represents the composite difference in the angles of the right and left eyes, i.e., the alignment, while the y-axis represents normalized cumulative sum probability. Red lines indicate when DBS was off, and blue lines represent STN-DBS on periods. The black line denotes the healthy control. According to STN-DBS effects the 12 PD participants were divided in three groups. Lack of STN-DBS effects was Group 1 (n=4). There was significant improvement with STN-DBS in Group 2 (n=6), while the Group 3 had worsening of strabismus with STN-DBS (n=2).

The next step of our analysis examined the mechanistic underpinnings of the varying STN DBS effects on PD participants. The results examined the hypothesis that the DBS of the dorsal STN improves the strabismus angle in PD. We examined the volume of tissue activation models in all PD participants, belonging to Group 1, Group 2, or Group 3 as noted above. The Group 2 PD participants who had robust strabismus which improved with STN DBS had the VTA located in the dorsal STN region, 3.0±0.8 mm dorsal to the center of STN (orange bar, **Fig 3A**). The Group 3 PD participants who had significant worsening of strabismus with STN DBS had the VTA located in the ventral STN, 0.7±1.6mm ventral to the center of STN (grey bar, **Fig 3A**). Other DBS VTA parameters, such as the medio-lateral or antero-posterior locations within the STN, or the VTA to STN overlap were not different between Group 2 and 3 (**Fig 3B-D**). The Group 1 PD participants who only had trendwise improvement with STN DBS had VTA parameters comparable to Group 2 (**Fig 3A-D**), however, they had minimal strabismus at baseline to show any improvement.

**Figure 3:**
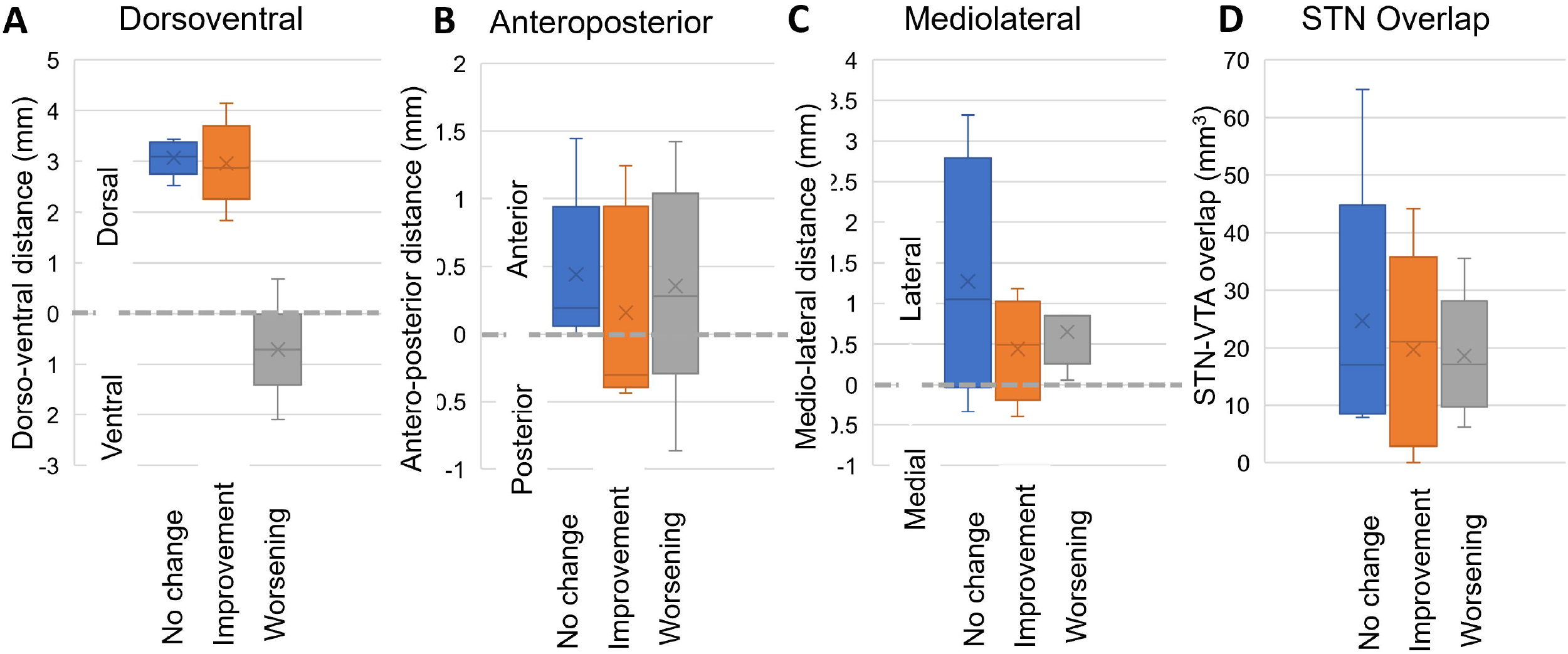
Location specific effects of STN-DBS on strabismus in PD. (A) Among three groups (unchanged, improved, and worsening), the improvement was associated with dorsal location of STN-DBS. The volume of activated tissue in ventral STN was associated with worsening of strabismus. (B,C,D) Other volume of tissue activation parameters, such as antero-posterior location (panel B), mediolateral location (panel C), and STN-volume of tissue activation overlap (panel D) did not determine the effects on strabismus.

In summary, this study investigated strabismus in PD and evaluated the effects of STN DBS on strabismus. Objective measures revealed varying strabismus severity in our PD participants. 33% of patients exhibited minimal misalignment unaffected by STN DBS. Among the remaining 66% with significant strabismus, 75% experienced improvement with STN DBS, while 25% saw worsening. All participants had improvement in PD motor symptoms. Patient-specific computational models of DBS VTA indicated that modulation of the dorsal STN correlated with strabismus improvement while maintaining its effects on PD motor symptoms, whereas modulation of the ventral STN was associated with deterioration. No significant differences were observed in other VTA parameters.

## Discussion

STN DBS is primarily used to treat motor symptoms of PD, such as tremors, rigidity, and bradykinesia, while improving the motor function and reducing the medication burden. Eye movement abnormalities affecting the simultaneous movements of both eyes and eye alignment, strabismus, are very common in PD [3-5, 7, 10, 16-18, 21]. This study examined whether STN DBS can improve strabismus in PD while preserving its effects on motor symptoms. We found that all PD participants had strabismus to a variable extent, and the STN DBS modulated strabismus in 66% of them. Among those who had a significant effect, 75% had improvement in strabismus angle, while 25% had worsening. The group with the lack of STN DBS effect had minimal strabismus at baseline, and their DBS activation parameters are indifferent compared to the group when there was a significant improvement. The group whose strabismus worsened with STN DBS had comparable volume of VTA as the group who had an improvement, but its location was ventral to the center of the STN.

Our results provide the following key messages:

1. Strabismus was observed in all PD participants, and it significantly affected 75% of the participants. Phoria was more robust in PD compared to tropia.
2. STN DBS changed strabismus when it was significant – improving it in two-thirds the PD cohort.
3. VTA in the dorsal STN improved strabismus, but VTA in the ventral STN worsened it.

These results are consistent with previous reports of highly prevalent strabismus leading to double vision in PD [4, 12, 13, 18, 29]. Worsening of monocular deviation, suggesting worse form of phoria, is also consistent with previous studies highlighting decreased foveation and convergence capabilities [12, 23, 30].

Our findings suggest that both dorsal and ventral STN are involved in the control of binocular alignment – VTA in dorsal STN improves the strabismus while ventral STN DBS worsens it. In other words, two parts of STN might modulate binocular control via distinct mechanisms. The results are further supported by well-known anatomical evidence from non-human primate studies. The ventral STN is known to have visuo-oculomotor neurons [36-40]. Eye movement sensitive neurons in the ventral STN receive input from the frontal and supplementary eye fields [41-43]. These regions then project to the superior colliculus via substantia nigra pars reticulate (SNr). The superior colliculus, via the nucleus reticularis tagmenti pontis, projects to the SOA – the midbrain region critical for binocular control [26, 27, 44]. In contrast, dorsal STN and zona incerta which are immediately dorsal to the dorsal STN are connected to the pre-cerebellar nuclei. The latter then projects to the deep cerebellar nuclei, and then to the SOA [42, 45, 46]. In other words, both ventral and dorsal STN can modulate binocular control putatively via involvement of the SOA. However, considering our findings, the physiological effects of the dorsal STN stimulation are opposite compared to ventral – the latter worsening the phoria and tropia in PD.

Our pilot investigation marks a pioneering effort, featuring several notable aspects. It stands as the inaugural study to quantify strabismus, phoria, and tropia in PD using advanced high-resolution oculography. Moreover, it delves into the impact of STN DBS on strabismus, employing a unique combination of objective oculographic evaluation and correlating structural volumes of STN tissue activation with DBS computational models.

It is important to acknowledge a critical limitation in the form of a modest sample size. Numerous challenges hindered the assembly of a larger participant pool necessary for robust statistical analyses. A primary obstacle was the requirement for a cohort devoid of significant tremor or dyskinesia, particularly when the DBS is turned off. Securing PD participants with minimal tremor at baseline, in STN DBS off state, posed a considerable challenge, as tremors (or dyskinesia), when present, could affect head movement and induce compensatory vestibulo-ocular reflex (VOR) during oculography. This “contaminant” in oculographic data could compromise the quality of recordings[47-51].

Another significant constraint involved identifying participants who were capable of successfully completing the experiment with the DBS turned off, without encountering notable cognitive or mental challenges. This additional factor further limited the pool available for assessment. Despite these challenges, we were able to include enough patients, resulting in consistent findings. Notably, our baseline assessment of strabismus aligns with clinical observations, demonstrating a correlation between objective measurements and subjective clinical impressions.

In conclusion, our study focused on assessing the impact of STN DBS on strabismus in PD. While STN DBS is commonly utilized for motor symptom management in PD, our findings revealed that all PD participants exhibited varying degrees of strabismus. STN DBS influenced strabismus in 66% of participants, with 75% experiencing improvement and 25% experiencing worsening. Notably, dorsal STN stimulation improved strabismus, whereas ventral STN stimulation worsened it. These results suggest distinct modulatory mechanisms in different parts of the STN for binocular control. Our study contributes to existing literature on strabismus prevalence in PD and enhances understanding of the physiological effects of dorsal and ventral STN stimulation on ocular functions. This multidisciplinary approach, integrating movement disorders neurology, visual neuroscience, and computational modeling of DBS, holds promise for developing effective treatments targeting visual abnormalities in PD and improving the overall well-being of affected individuals.

## Data Availability

All data produced in the present study are available upon reasonable request to the authors, after completing necessary documentation with the department of Veterans Affairs.

## Funding and acknowledgements

Shaikh was supported by the Career Development Grant from the American Academy of Neurology, George C. Cotzias Memorial Fellowship, Network Models in Dystonia grant from the Dystonia Medical Research Foundation, Department of VA Rehab R&D Spire (I21RX003878), CareSource Ohio Community Partnership Grant, and philanthropic funds for the Department of Neurology at University Hospitals. Shaikh holds Penni and Stephen Weinberg Chair in Brain Health. This work was also supported by the National Institutes of Health (NIH) grant support to McIntyre NIH R01NS119520.

